# Real-world data validation of the PurIST pancreatic ductal adenocarcinoma gene expression classifier and its prognostic implications

**DOI:** 10.1101/2023.02.23.23286356

**Authors:** Stephane Wenric, James M. Davison, John Guittar, Gregory M. Mayhew, Kirk D. Beebe, Yun E. Wang, Amrita A. Iyer, Hyunseok P. Kang, Michael V. Milburn, Vincent Chung, Tanios Bekaii-Saab, Charles M. Perou

## Abstract

**Background:** Pancreatic ductal adenocarcinoma (PDAC) is amongst the deadliest cancers, with few modern tools to inform patient prognosis and help guide treatment options. Transcriptome-based molecular subtyping is one emerging technology that has been employed to help patients optimize available therapeutic approaches. Here we retrospectively demonstrate the clinical validity of PurIST (Purity Independent Subtyping of Tumors), an RNA-based classifier that divides PDAC patients into two subtypes with differential prognoses, as a validated laboratory-developed test (LDT) on the Tempus Labs sequencing platform.

**Methods:** A cohort comprising 258 late-stage PDAC patients with available transcriptomic and outcomes data was drawn from the Tempus clinicogenomic database and classified using PurIST into one of two subtypes (“Basal” or “Classical”). Differences in patient survival from the date of diagnosis were compared between subtypes, and between two common first-line treatment regimens, FOLFIRINOX, and gemcitabine + nab-paclitaxel.

**Results:** Of the 258 PDAC patients in the validation cohort, PurIST classified 173 as classical subtype, 59 as basal subtype, and 26 as no-calls. Reinforcing previous findings, patients of the basal subtype had significantly lower overall survival than those of the classical subtype. Notably, differential survival by subtype was significant among the subset of patients on FOLFIRINOX, but not those on gemcitabine + nab-paclitaxel.

**Conclusions:** The implementation of PurIST on a high-throughput clinical laboratory RNA-Seq platform and the demonstration of the model’s clinical utility in a real-world cohort together show that PurIST can be used at scale to refine PDAC prognosis and thereby inform treatment selection to improve outcomes for advanced-stage PDAC patients.

## Introduction

Pancreatic cancer represents approximately 3.2% of all new cancer diagnoses with an estimated 60,430 new cases diagnosed in 2021 (Park et al. 2012, SEER Cancer Stat Facts), while pancreatic ductal adenocarcinoma (PDAC) constitutes the most prevalent type of pancreatic neoplasm, accounting for more than 90% of all pancreatic cancer cases. To date, PDAC is the fourth most frequent cause of cancer-related deaths worldwide with a 5-year overall survival below 8% (Orth et al. 2019; Sarantis et al. 2020). When known tumor-specific alterations are detected in PDAC patients, targeted treatment can be used to significantly improve progression-free and overall survival (Lowery et al. 2017; Aguirre et al. 2018). Unfortunately, only 26% of pancreatic cancer patients have these types of actionable molecular alterations (Pishvaian et al. 2020), highlighting the need for a more general approach to optimizing treatment selection.

Transcriptome-based molecular profiling of tumor tissue can offer additional insight into tumor biology and help guide treatment decisions. For example, transcriptomic data can be used to detect gene expression networks or the activity of oncogenic pathways and support the development of more targeted therapies (Malone et al 2020). Gene expression data can also be used to define and identify cancer subtypes that differ in their rates of disease progression or responses to specific therapies (Weigelt et al. 2009; Kim et al. 2019). Already, transcriptome-based molecular profiling has been used to predict the risk of recurrence in prostate (Long et al. 2014), breast (Wallden et al. 2015), and lung cancers (Sun et al. 2021; Ma et al. 2021), and to guide treatment decisions in lung, breast, and bladder cancers (Robinson et al. 2017; Benayed et al. 2019; Adashek et al. 2020; Wise and Solit 2019; Rodon et al. 2019).

Several transcriptome-based schemas have been proposed to enable molecular subtyping of PDAC (Collisson et al. 2011; Bailey et al. 2016; Puleo et al. 2018; Moffitt et al. 2015), with the Moffitt et al. schema showing strong prognostic value and performing well independently of biopsy purity (Rashid et al. 2020; TCGA 2017). Moffitt et al. used the expression of tumor intrinsic genes to identify two PDAC subtypes, “basal-like” (hereafter “basal”) and “classical,” which remain stable and distinct regardless of patient treatment experience (Moffitt et al. 2015). Rashid et al. (2020) formalized the Moffitt schema with Purity Independent Subtyping of Tumors (PurIST), a classifier with intra-sample normalization to enable single-sample subtyping suitable for a clinical setting. PurIST consistently recapitulated Moffitt basal and classical subtype calls with high fidelity across multiple platforms (microarray, NanoString NGS, and Illumina NGS) and sample types (flash-frozen, FFPE), and on publicly available datasets (TCGA PDAC) (Rashid et al. 2020). Importantly, preliminary evidence suggests that PurIST subtypes may have prognostic value, with classical patients showing significantly higher response rates and survival benefits from leucovorin-fluorouracil-irinotecan-oxaliplatin (FOLFIRINOX) than basal patients (O’Kane et al. 2020; Rashid et al. 2020). PurIST is currently being further evaluated for treatment matching in resectable and borderline-resectable PDAC patients in a phase II trial (NCT04683315).

In this study, we report the results of a prospectively designed retrospective clinical validation of the PurIST classifier as a laboratory-developed test (LDT) on the Tempus Labs sequencing platform. We validate PurIST using a real-world dataset of 258 patients with advanced PDAC who received one of the two most-administered first-line treatments, FOLFIRINOX or gemcitabine + nab-paclitaxel. All samples were processed on the Tempus xT RNA sequencing whole exome capture transcriptome assay. The implementation of PurIST on a high-throughput clinical laboratory RNA-Seq platform and the demonstration of the model’s clinical utility in a real-world cohort together show that PurIST can be used at scale to refine PDAC prognosis and thereby inform treatment selection and improve outcomes for advanced-stage PDAC patients.

## Methods

### Sample Selection

Three PDAC cohorts were drawn from the Tempus clinicogenomic database, one for analytical validation of PurIST at Tempus, one to test for concordance across platforms, and the third for clinical validation. For the analytical validation (N = 36 reference samples), patients with any stage of PDAC were selected. For the platform concordance cohort (N=100), we selected patients with early-stage PDAC to maximize similarity to the original Moffitt et al. (2015) cohort; namely, those with Stage I-III resectable tumors and biopsies exclusively from primary tumors in the pancreas. For the validation cohort (N=258), we selected patients with late-stage PDAC, defined as Stage III-IV unresectable tumors, and biopsies from either primary or metastasized tumors, to better reflect the types of patients that will be seeking PurIST results in a clinical setting. Additionally, patients in the platform concordance and clinical validation cohorts had no prior resections, were treatment naive at the time of biopsy, were treated with either FOLFIRINOX or gemcitabine + nab-paclitaxel as first-line (1L) systemic therapy, had biopsy sequencing performed no more than 30 days prior to 1L treatment start date, and had available outcomes data. Patient data were de-identified according to HIPAA guidelines. Line of therapy was determined using clinician notes and clinical abstraction methods. Dates of diagnosis and sequencing spanned years 2009 – 2021 and 2017 - 2022 for the two cohorts, respectively. Clinical abstraction included records that had data for pancreatic cancer diagnosis date, age, gender, race, status (primary vs metastatic), stage (resectable vs non-resectable). Tempus xT sequencing was completed between 2017 - 2022. The study design, including analyses and endpoints, was prospectively defined before development of the validation cohort.

### Clinical Data Abstraction

Clinical data were extracted from the Tempus real-world oncology database. This encompassed longitudinal structured and unstructured data from geographically diverse oncology practices, including integrated delivery networks, academic institutions, and community practices. Records included in this study may have been obtained in partnership with ASCO CancerLinQ. Structured data from electronic health record systems were integrated with unstructured data collected from patient records via technology-enabled chart abstraction and corresponding molecular data, if applicable. Data were harmonized and normalized to standard terminologies from MedDRA, NCBI, NCIt, NCIm, RxNorm, and SNOMED. Additional mortality data were obtained from Datavant, an organization that augments Social Security Administration death master files with information from newspapers, funeral homes, and memorials to construct an individual-level database of more than 80% of US deaths annually (Datavant, Mortality Data in Healthcare Analytics, 2021). Patients with no recorded date of death across all mortality sources were censored at the date of last recorded interaction with the medical system (i.e., date of last follow-up).

### DNA and RNA sequencing

De-identified records from 394 Formalin-Fixed Paraffin-Embedded (FFPE) tumor samples from PDAC with DNA and RNA sequencing data were selected from the Tempus Oncology Database. All samples underwent DNA and RNA sequencing using the Tempus xT targeted panel. Tempus xT is a next-generation sequencing assay employed for the detection of specific cancer targets by sequencing tumor samples from patients (saliva or blood samples) when available. Tempus xT detects insertions and/or deletions, single-nucleotide variants, and copy number variants in 648 genes (spanning ∼3.6Mb long genomic region) along with chromosomal rearrangements in 22 genes with high specificity and sensitivity. The assay also evaluates transcriptomic alterations from whole-transcriptome RNA sequencing data, including variations in gene expression, altered splicing, and fusions. Sample preparation, DNA sequencing, and RNA sequencing were conducted as described (Leibowitz et al. 2022; Michuda et al. 2022).

### The PurIST model

PurIST is a Top Scoring Pairs (k-TSPs) gene signature model that assesses rank-based expression levels of 16 genes sorted into 8 gene pairs from a single sample to estimate the likelihood of that sample being basal (Rashid et al. 2020).

### Robustness of the PurIST model on the Tempus platform

To confirm that PurIST performance was not sensitive to differences in sequence processing platforms, we compared results from samples bioinformatically processed according to the pipeline described in Rashid et al. to those processed as part of the whole-transcriptome xT assay at Tempus. This comparison included a difference in normalization method, namely transcripts per million (Rashid et al.) vs. the Tempus xT method of normalization based on transcript length, GC content, and library size as described in Leibowitz et al. (2022). Similarly, to confirm that PurIST performance was not sensitive to differences in model training datasets, we compared results from the original model as described by Rashid et al. to those from a new version of the model retrained on Tempus transcriptomic data. More specifically, the model training process described in Rashid et al. (2020) was used to generate new Tempus-specific PurIST gene-pair coefficients, and intercepts and subtype classifications from both model versions were compared for samples from the platform concordance cohort (N=100).

Analytical validation of PurIST on the Tempus platform comprised four stages: limit of detection (LOD), intra-assay precision, inter-assay precision, and inter-sequencer concordance. To establish the lower limit of detection in terms of tumor purity, four tumor samples with at least 50% purity were diluted to 20%, 30%, and 40% purity using unmatched adjacent normal tissue obtained from patients with pancreatic cancer. Diluted samples were sequenced and analyzed in triplicate at all three dilution levels. Intra-assay precision was determined by processing eight samples in triplicate on a single sequencing run. Inter-assay precision was determined by processing 12 samples in triplicate by at least two different operators on different equipment, on different days, and using different reagent lots. Inter-instrument concordance was determined by sequencing nine samples in triplicate across three different Illumina NovaSeq sequencers. All stages of the analytical validation were performed at Tempus Labs in Chicago, IL; a second analytical validation was performed in parallel at Tempus Labs in Durham, NC.

### Confidence scoring and implementation of a no-call zone

Like any classifier that uses continuous measures to group patients into discrete bins, PurIST can give low-confidence or indeterminate results when the expressions of gene pairs used in the algorithm provide results that are within statistical margins of error between those defining the classical and basal subtypes. While it is not important to account for such indeterminate samples in population-level analyses when mean effects are the primary concern, it is important to flag them when reporting LDT results to individual patients to ensure a high degree of patient-level reproducibility. Hence, we developed a simulation-based approach to calculate confidence scores for each sample by evaluating across all gene pairs the proximity of the paired values, the subsequent likelihood of rank inversion, and subtype flipping as a result. Samples with confidence scores below 0.85 were flagged as no-calls and were censored from analytical validations.

Confidence scores were generated for each sample as follows:

1. For each observed expression value for each PurIST gene, a predicted standard deviation was estimated using a simple linear model:

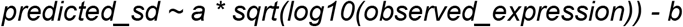
2. where constants *a* = −0.08344563 and *b* = 0.2120142 as determined by fitting the model to a dataset comprising 1000 replicate runs of a single universal human reference sample on the Tempus RNA-Seq pipeline. The linear model predicts standard deviation based on mean expression alone and is agnostic to gene identity.
3. A new set of 16 gene log-transformed expression values was drawn from 16 normal distributions defined using the observed log-transformed expression values as their means along with their respective predicted standard deviations. The PurIST algorithm was then re-applied to the new set of simulated expression values to determine the predicted classification.
4. The process in (2) was repeated 10000 times, and the frequencies of basal and classical classification outcomes were tabulated.
5. Confidence score was calculated as the number of times the majority classification was observed divided by the total number of simulations (10000); as such, scores ranged from 0.5 (very low confidence; equivalent to a coin flip) to 1.0 (very high confidence; zero discordance expected).

### Validation of the PurIST model in a RWE PDAC cohort

Kaplan-Meier analysis of overall survival (OS) from the date of diagnosis was compared between subtypes and other cohort groupings and assessed with log-rank tests. Univariable and multivariable statistical associations to OS were determined using Cox proportional hazards (cox PH) models. The 12-month survival rates were compared between groups using two-proportion z-tests.

### Clinical and molecular associations with PurIST subtypes and outcome measures

Associations between clinical or molecular variables and OS were assessed using the same methods as above. A comparison of gene-level mutation burden was performed using mutations deemed pathogenic and likely pathogenic as per the Tempus Knowledge Database (KDB) (Beaubier et al. 2019). The proportion of samples harboring gene-specific mutations and proportions of specific amino acid changes were compared using Fisher’s exact tests. Differential gene expression between subtypes was determined using the Wilcoxon test. Cox PH models were used to examine gene expression associations with OS.

## Results

To begin our PDAC tumor subtyping studies, qualifying Tempus PDAC samples (N=358) were grouped into a platform concordance cohort (N=100), consisting of patients with early-stage resectable tumors, and a clinical validation cohort (N=258), consisting of patients with late-stage unresectable tumors. The overall schema of the study is shown in Figure 1, and the clinical and demographic characteristics of both cohorts are shown in the Supplementary Materials section and Table 1, respectively.

**Figure 1.**
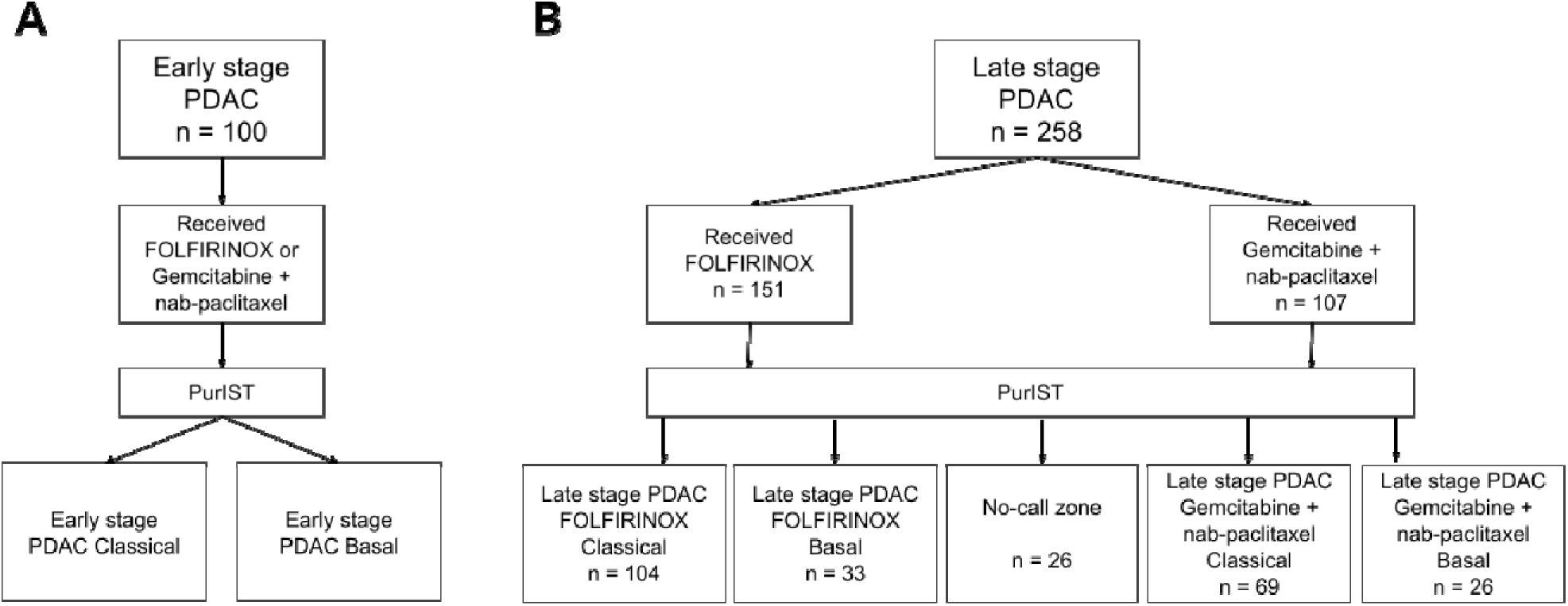
Study workflow. **A**. Platform concordance cohort. **B**. Retrospective clinical validation cohort.

**Table 1.** Clinical validation cohort.

| Characteristic | Basal, N = 59<sup>1</sup> | Classical, N = 173<sup>1</sup> | p-value<sup>2</sup> |
| --- | --- | --- | --- |
| **Gender** |  |  | 0.003 |
| Female | 15 (25%) | 82 (47%) |  |
| Male | 44 (75%) | 91 (53%) |  |
| **Age at diagnosis** | 66 (61, 70) | 66 (58, 72) | 0.7 |
| **DDR mutation** | 1 (1.7%) | 16 (9.4%) | 0.079 |
| Unknown | 1 | 3 |  |
| **regimen** |  |  | 0.6 |
| FOLFIRINOX | 33 (56%) | 104 (60%) |  |
| gemcitabine + paclitaxel | 26 (44%) | 69 (40%) |  |
| **Stage** |  |  | >0.9 |
| Stage 3 | 3 (5.1%) | 9 (5.2%) |  |
| Stage 4 | 56 (95%) | 164 (95%) |  |
| **Histology** |  |  | 0.5 |
| Adenocarcinoma | 36 (63%) | 104 (62%) |  |
| Adenocarcinoma, metastatic | 15 (26%) | 48 (29%) |  |
| Carcinoma, metastatic | 1 (1.8%) | 0 (0%) |  |
| Infiltrating duct carcinoma | 5 (8.8%) | 15 (9.0%) |  |
| Unknown | 2 | 6 |  |
| **Liver metastasis** | 46 (78%) | 119 (69%) | 0.2 |
| **ECOG** |  |  | 0.4 |
| 0 | 4 (33%) | 32 (48%) |  |
| 1 | 6 (50%) | 29 (43%) |  |
| 2 | 1 (8.3%) | 3 (4.5%) |  |
| 3 | 1 (8.3%) | 3 (4.5%) |  |
| Unknown | 47 | 106 |  |
| <sup>1</sup>n (%); Median (IQR) |  |  |  |
| <sup>2</sup>Pearson's Chi-squared test; Wilcoxon rank sum test; Fisher's exact test |  |  |  |

**Table 2.**
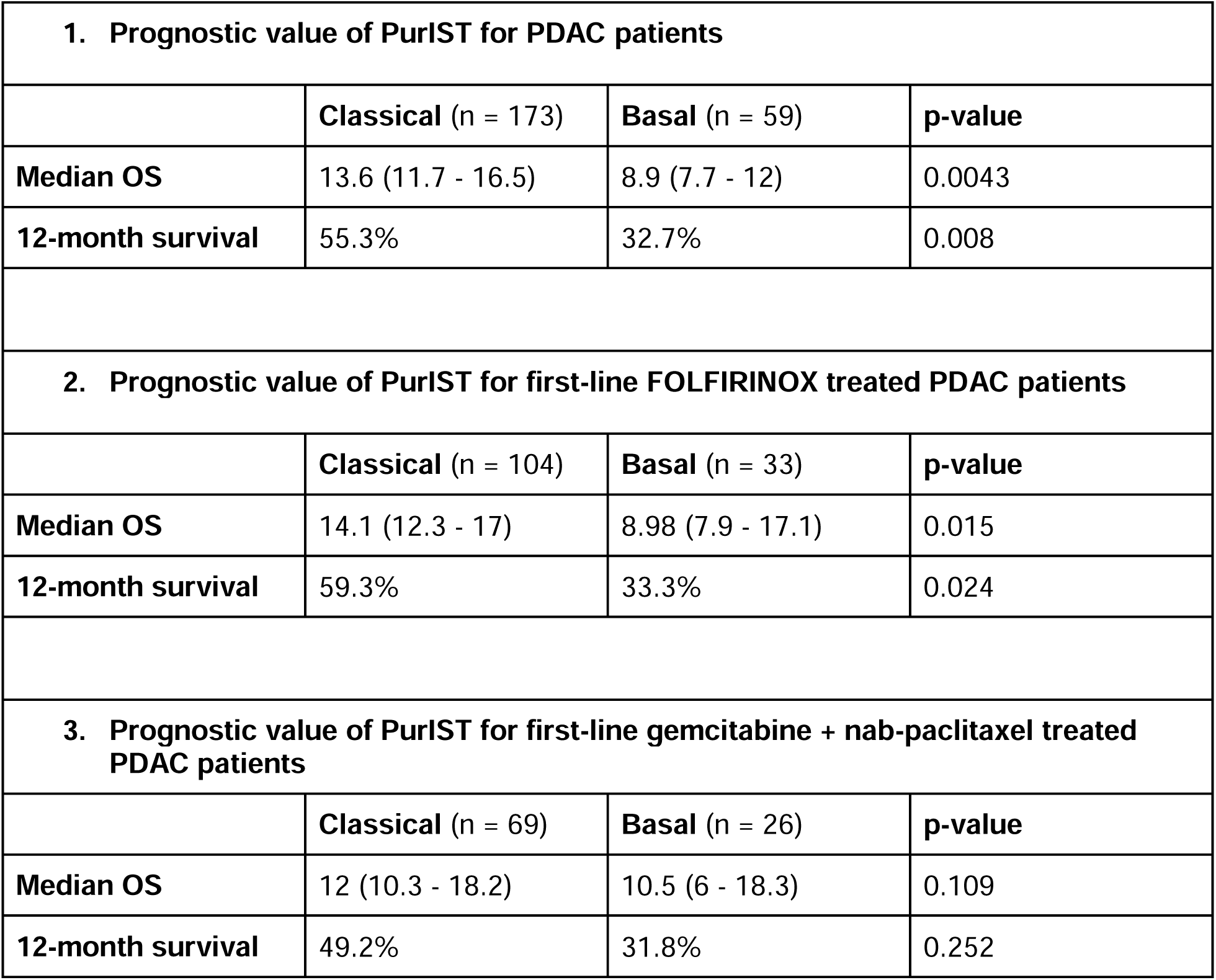
Validation of the PurIST model in a RWE PDAC cohort. 1. Median OS and 12-month survival of the PurIST subtypes ascertained in the full cohort (n = 232). 2. Median OS and 12-month survival of the PurIST subtypes ascertained in the FOLFIRINOX treatment group (n = 137). 3. Median OS and 12-month survival of the PurIST subtypes ascertained in the gemcitabine + nab-paclitaxel treatment group (n = 95).

### Robustness of the PurIST model on the Tempus platform

Samples processed using both Rashid et al. and Tempus bioinformatics pipelines (N = 100) were 98% concordant in their PurIST subtypes, suggesting that the classifier is robust to differences among sequencing data processing pipelines (Supplementary Figure 1). Likewise, the original PurIST model (Rashid et al. 2020) and the model retrained using data from the Tempus assay were 98% concordant, suggesting that the classifier is robust to training set differences. Given that the two versions of the PurIST model performed equivalently, the original PurIST model as described in (Rashid et al. 2020) was used to subtype patients in the validation cohort.

Results from the four stages of the analytical validation showed 100% agreement within all sets of replicates, indicating that the PurIST algorithm generates reliable and reproducible subtype classifications on the Tempus sequencing pipeline. Of the 36 reference samples initially selected for the analytical validation, two were identified as no-calls and removed. The remaining 34 reference samples (15 basal, 19 classical) were diluted with normal tissue where applicable, subsampled in triplicate, and submitted for sequencing, resulting in 138 attempted replicates. Of these, eight failed sequencing, resulting in 130 successfully processed replicates. Together, the four stages of the analytical validation show that (1) PurIST subtypes can be reliably determined at tumor purities as low as 20%, (2) subtyping results are consistent when run in parallel on the same sequencing run, (3) subtyping results are reproducible even when processed by different users on different equipment, and (4) subtyping results are not sensitive to minor differences between Illumina NovaSeq sequencers. For specific sample sizes and additional details about the analytical validation, refer to Supplementary Table 2.

### Validation of the PurIST model in a RWE PDAC cohort

#### Patient validation cohort

Of the 258 patients in the validation cohort, 26 were determined to be no-calls and dropped from the analysis. Of the remaining 232 patients, PurIST classified 173 (74.6%) tumors as the classical subtype and the remaining 59 (25.4%) as the basal subtype (Table 1). Basal patients were significantly more likely than classical patients to be male (75% vs. 53%, respectively; p = 0.003). Basal patients were also more likely than classical patients to have a liver metastasis (78% vs. 69%), and less likely than classical patients to have a pathogenic or likely pathogenic mutation in a DNA Damage Repair (DDR) gene (1.7% vs 9.4%) defined as *BRCA1, BRCA2, PALB2, ATM, ATR*, ATRX, *BAP1, BARD1, BRIP1, CHEK1, CHEK2, RAD50, RAD51, RAD51B, FANCA, FANCC, FANCD2, FANCE, FANCF, FANCG, FANCL*, but these associations were not significant. Following the current standard of care, patients who were younger and had lower ECOG scores were more likely to receive FOLFIRINOX than gemcitabine + nab-paclitaxel.

#### The prognostic value of PurIST for PDAC patients

PurIST subtype classification was broadly prognostic of survival for patients in the validation cohort. Specifically, basal patients had significantly shorter OS than classical patients (HR = 1.61; 95% CI: 1.16-2.25; p = 0.0043), with median OS values of 8.93 months (95% CI: 7.73-12.03) and 13.6 months (95% CI: 11.67-16.5), respectively (Figure 2A). Similarly, basal patients were significantly less likely than classical patients to survive to 12 months (32.7% vs. 55.3%, respectively; p = 0.008), recapitulating the prognostic subtype association observed in prior studies (Moffitt et al. 2015; Rashid et al. 2020).

**Figure 2.**
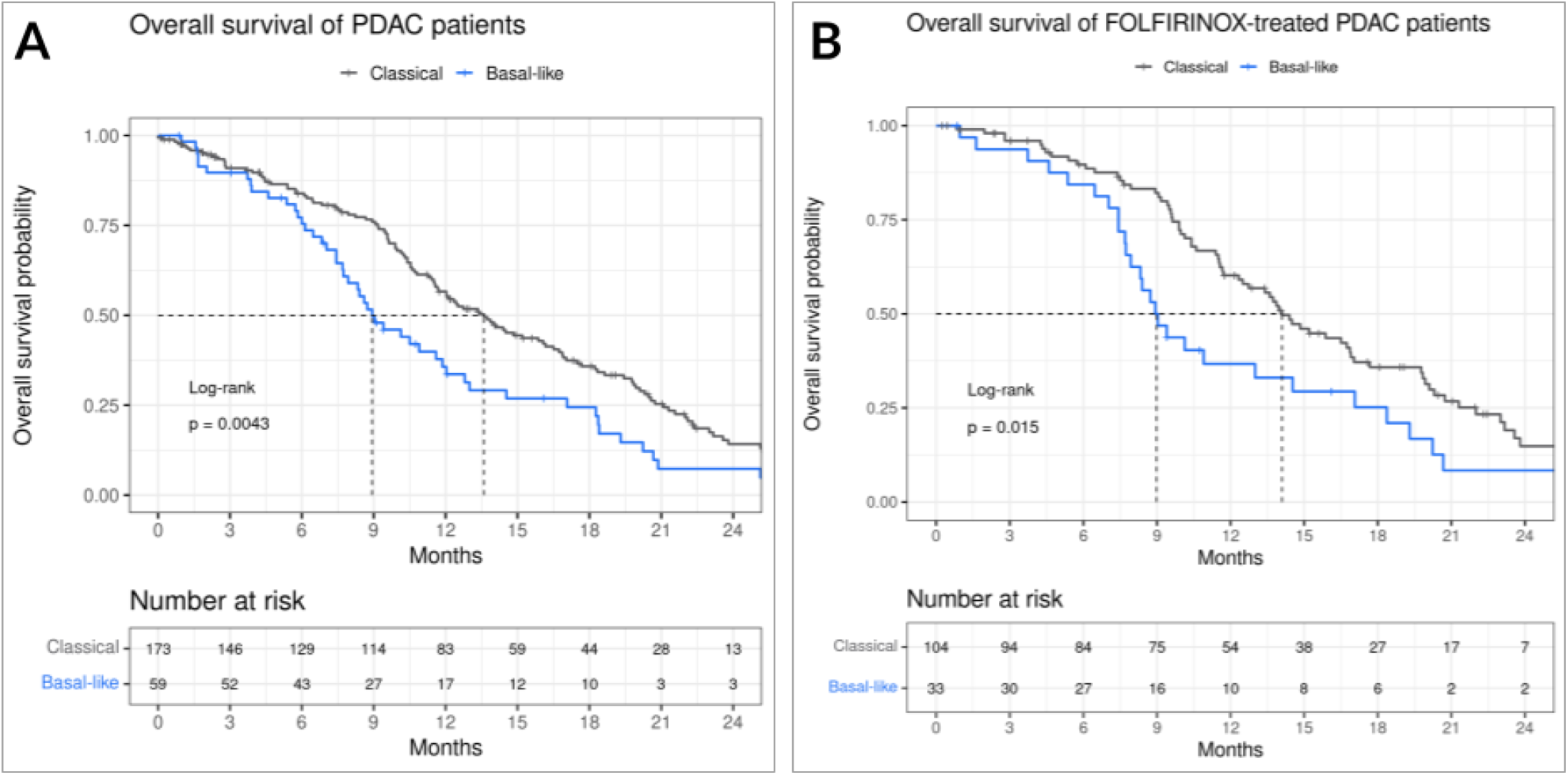
Validation of the PurIST model in a RWE PDAC cohort. **A**. Overall survival of the full cohort (n = 232). **B**. Overall survival of the FOLFIRINOX treatment group (n = 137).

#### Prognostic value of PurIST for first-line FOLFIRINOX treated PDAC patients

PurIST subtype classification was significantly prognostic in the subset of patients who received FOLFIRINOX as 1L, but not in the subset of patients who received gemcitabine + nab-paclitaxel as 1L. Among patients treated with FOLFIRINOX (n = 137), basal patients (n = 33) had significantly shorter OS than classical patients (n = 104) (HR = 1.7; 95% CI, 1.1-2.63; p = 0.015), with median OS values of 8.98 months (95% CI: 7.93-17.07) and 14.1 months (95% CI: 12.33-17.03), respectively (Figure 2B). In addition, FOLFIRINOX-treated basal patients were significantly less likely than classical patients to survive to 12 months (33.3% vs. 59.34%, respectively; p = 0.024). Among patients treated with gemcitabine + nab-paclitaxel (n = 95), basal patients (n = 26) did not differ significantly from classical patients (n = 69) in their median OS (basal = 10.5 months, 95% CI: 6-18.3 vs; classical= 12 months, 95% CI: 10.3-18.2; p = 0.1087; Supplementary Figure 3) or 12-month survival (basal = 31.8%, classical = 49.2%; chi-squared p-value = 0.25).

### Clinical and molecular associations with PurIST subtypes and outcome measures

#### Clinical variables associated with prognosis

We next assessed other relevant clinical variables that were potentially predictive of overall survival in this cohort. We found sex to be highly prognostic for PDAC patients (HR = 1.63, 95% CI = 1.2-2.21, p = 0.0014), irrespective of 1L therapy administered, with males having a worse prognosis both overall and within subtypes and treatment groups. Importantly, PurIST subtype remained a significant predictor of OS in a multivariable Cox PH model accounting for sex (HR = 1.44, CI = 1.02-2.02, p = 0.038). However, in a similar multivariable model including sex and the PurIST subtype as covariates, the PurIST subtype was only marginally associated with survival among FOLFIRINOX-treated patients (HR = 1.53, CI = 0.96-2.43, p = 0.074). The presence of liver metastases was marginally associated with survival, but only among patients treated with FOLFIRINOX (HR = 1.57, CI = 0.98-2.52, p = 0.057), not among patients treated with gemcitabine + nab-paclitaxel. Within the subgroup of patients with liver metastases, the PurIST subtype was significantly associated with survival both for the FOLFIRINOX-treated group and for the two regimens together (HR = 2.02, CI = 1.22-3.35, p = 0.0054 and HR = 1.64, CI = 1.12-2.41, p = 0.011 respectively), as basal patients with liver metastasis had a worse prognosis than classical patients with metastasis. The presence of DNA Damage Repair (DDR) mutations was significantly associated with survival in the full cohort in a univariate analysis (HR = 0.52, CI = 0.28-0.96, p = 0.034), however in a multivariable Cox PH model including the PurIST subtype, only the PurIST subtype was significantly associated with survival, both in the FOLFIRINOX treated cohort and in the combined regimens cohort (Supplementary Figure 4). In a univariate model, neither age nor histological site was associated with prognosis. In a multivariable Cox PH analysis including sex, age at diagnosis, histological site, the presence of liver metastasis, and of DDR mutations, and ECOG score the PurIST subtype remained significantly associated with OS (HR = 6.26, 95% CI = 1.49-26.38, p = 0.012) in the FOLFIRINOX-treated group (Figure 3A).

**Figure 3.**
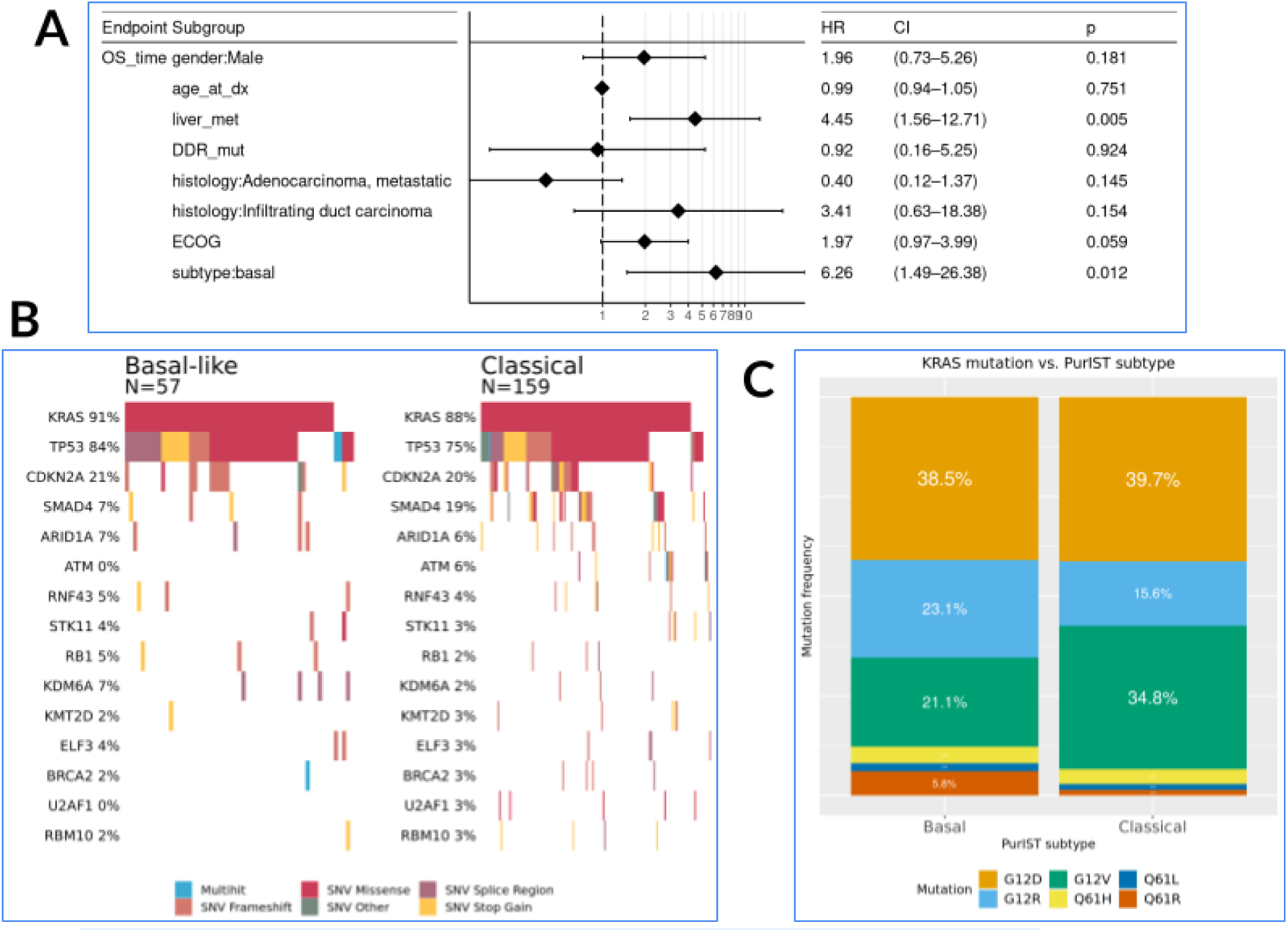
**A**. Multivariable survival analysis including clinical covariates in the FOLFIRINOX treatment group (n = 41 patients with recorded ECOG score, due to missingness). **B**. Comparative mutation rates for basal and classical cohorts with available DNA samples. *SMAD4* and *ATM* mutations are more common in classical patients than in basal patients (19% vs. 7%; p = 0.05; 6% vs. 0%, p = 0.07 respectively). **C**. Comparative KRAS codon mutation distributions between basal and classical patients.

#### KRAS coding mutations and other commonly mutated genes

Previous studies have shown that select gene alterations can influence PDAC disease status in both classical and basal patients (Saiki et al. 2021). We tested for differences in mutation rates, gene expression, and amino acid changes using Fisher’s tests in *KRAS, TP53, CDKN2A*, and *SMAD4*, four genes previously associated with PDAC (Cicenas et al. 2017), as well as in 10 additional genes with high mutation prevalence in the validation cohort (Figure 3B). Single nucleotide variants for *KRAS, TP53*, and *CDKN2A* were similarly prevalent in patients from both PurIST subtypes, while *SMAD4* and *ATM* mutations were more common in classical patients than in basal patients (19% vs. 7%; p = 0.05; 6% vs. 0%, p = 0.07 respectively), see Figure 3B). Gene expression in *KRAS, TP53*, and *SMAD4* did not differ significantly between the two subtypes, while gene expression in *CDKN2A* was significantly higher in basal patients (p = 0.031). The frequency of *KRAS* codon mutations differed nominally by PurIST subtype (p = 0.08), with G12V being more prevalent in classical patients (34.8% vs 21.1%) and G12R in basal patients (23.1% vs 15.6%), but these codons were not associated with differences in survival in a subtype-stratified analysis (Figure 3C). Our findings corroborate previous work highlighting the potential implications of codon 12-specific *KRAS* mutations in PDAC (Mueller et al. 2018; Smit et al. 1988). Additional Fisher tests on genes with high mutation prevalence did not yield a significant result.

## Discussion

In this study, we demonstrate the clinical validity of the Tempus PurIST molecular subtyping assay as a prognostic tool for PDAC patients when performed as an LDT on the Tempus xT sequencing platform. Validation was established using a predefined statistical plan, using an independent, retrospective, real-world cohort of advanced-stage (Stage III unresectable and Stage IV) PDAC patients undergoing biopsy before receiving first-line systemic therapy. While we recapitulated the results from previous studies (Moffitt et al. 2015; Rashid et al. 2020) in demonstrating that basal subtype patients have overall worse survival than classical subtype, we further extended the analysis and validated PurIST’s regimen-specific prognostic value. Amongst patients receiving 1L FOLFIRINOX, basal subtype patients had a significantly worse prognosis compared to classical subtype patients. This supplements the results of Rashid et al. to a survival endpoint and confirms that basal subtype patients not only have poor response rates to FOLFIRINOX but also poorer survival when being administered FOLFIRINOX.

In a previous study analyzing the same cohort, which did not require the implementation of a no-call zone, classical subtype patients receiving FOLFIRINOX as first-line therapy survived significantly longer than classical subtype patients receiving gemcitabine + nab-paclitaxel as first-line therapy (p = 0.046), while there was no significant difference in survival by regimen among basal subtype patients (Wenric et al. 2022). Implementation of a no-call zone in the clinical implementation of PurIST effectively reduced the number of samples included in the analysis presented here, resulting in a survival-based analysis that was directionally similar, but not statistically significant.

Given the real-world, retrospective nature of the validation cohort, some clinical variables were not balanced between treatment groups, with FOLFIRINOX-treated patients unsurprisingly being younger on average and having lower ECOG PS than the gemcitabine + nab-paclitaxel treated cohort. As a result, while directionally encouraging, future prospective studies should be conducted to confirm and further investigate the results described here. To the best of our knowledge, these findings represent the first clinical validation of PurIST as a molecular subtyping classifier providing FOLFIRINOX-specific prognostic value.

Clinical covariate analysis assessing the whole clinical validation cohort and within subtypes and treatment groups revealed that males have significantly worse overall survival both overall and within subtypes and treatment groups. This result aligns with findings from prior studies that males diagnosed with PDAC tend to have a worse prognosis than females (Jinkook Kim et al. 2021; Pijnappel et al. 2022). Additionally, patients with liver metastases had worse overall survival, both within the FOLFIRINOX treatment group (HR = 1.57, p = 0.057) and the gemcitabine + nab-paclitaxel treatment group (HR = 1.33, p = 0.27); similar results have also been observed by others (Suenaga et al. 2014; Groot et al. 2018). Finally, both the male sex and the presence of liver metastases were significantly enriched in basal subtype patients. This enrichment may be due to intrinsic differences in molecular drivers of disease, differences which might be molecularly recapitulated by the PurIST subtypes. Other studies have demonstrated that differential molecular drivers of disease aggressiveness are associated with the male sex (Natale et al. 2020; Hermann et al. 2021) and liver metastases (Paik et al. 2012; Sperti et al. 1997; Yachida et al. 2012), both of which are associated with the poorer prognosis basal subtype in our validation cohort. Importantly, PurIST remains significantly associated with survival even after accounting for these variables, confirming that PurIST has prognostic value and clinical utility in a heterogeneous demographic background.

In summary, these results, using the largest retrospective real-world PDAC cohort with RNA-Seq reported to date, demonstrate the clinical validity of the PurIST molecular subtyping classifier as a prognostic marker for PDAC patients receiving FOLFIRINOX when performed as a laboratory-developed test on the Tempus xT platform. As part of the comprehensive DNA and RNA profiling offered by the platform, including driver mutation status (e.g., *EGFR, NTRK, KRAS, TP53*) and HRD status, PurIST may help decision-making in the critical first-line setting and contribute to personalized care for PDAC patients.

## Data Availability

All data produced in the present work are contained in the manuscript.

## Notes

### Competing Interest Statement

Stephane Wenric, John Guittar, Yun E. Wang, Amrita A. Iyer, Hyunseok P. Kang were employed by Tempus Labs at the time of their contribution to the manuscript.
James M. Davison, Gregory M. Mayhew, Kirk D. Beebe, Michael V. Milburn were employed by GeneCentric at the time of their contribution to the manuscript.
Dr. Charles Perou is a board of directors member, equity stock holder, and consultant for GeneCentric.

### Funding Statement

This study did not receive any funding.

### Author Declarations

The study was performed using de-identified data and covered by an exempt determination from Advarra, Inc Institutional Review Board (IRB), Pro00042950

### Summary of Updates

Figure 3 and Table 1 revised.

## References

Adashek, Jacob J., Shumei Kato, Rahul Parulkar, Christopher W. Szeto, J. Zachary Sanborn, Charles J. Vaske, Stephen C. Benz, Sandeep K. Reddy, and Razelle Kurzrock. 2020. “Transcriptomic Silencing as a Potential Mechanism of Treatment Resistance.” JCI Insight 5 (11). https://doi.org/10.1172/jci.insight.134824.

Aguirre, Andrew J., Jonathan A. Nowak, Nicholas D. Camarda, Richard A. Moffitt, Arezou A. Ghazani, Mehlika Hazar-Rethinam, Srivatsan Raghavan, et al. 2018. “Real-Time Genomic Characterization of Advanced Pancreatic Cancer to Enable Precision Medicine.” Cancer Discovery 8 (9): 1096–1111.

Bailey, Peter, David K. Chang, Katia Nones, Amber L. Johns, Ann-Marie Patch, Marie-Claude Gingras, David K. Miller, et al. 2016. “Genomic Analyses Identify Molecular Subtypes of Pancreatic Cancer.” Nature 531 (7592): 47–52.

Benayed, Ryma, Michael Offin, Kerry Mullaney, Purvil Sukhadia, Kelly Rios, Patrice Desmeules, Ryan Ptashkin, et al. 2019. “High Yield of RNA Sequencing for Targetable Kinase Fusions in Lung Adenocarcinomas with No Mitogenic Driver Alteration Detected by DNA Sequencing and Low Tumor Mutation Burden.” Clinical Cancer Research: An Official Journal of the American Association for Cancer Research 25 (15): 4712–22.

Britta Weigelt, Frederick L Baehner, Jorge S Reis-Filho. 2009. “The Contribution of Gene Expression Profiling to Breast Cancer Classification, Prognostication and Prediction: A Retrospective of the Last Decade.” The Journal of Pathology, November. https://doi.org/10.1002/path.2648.

Collisson, Eric A., Anguraj Sadanandam, Peter Olson, William J. Gibb, Morgan Truitt, Shenda Gu, Janine Cooc, et al. 2011. “Subtypes of Pancreatic Ductal Adenocarcinoma and Their Differing Responses to Therapy.” Nature Medicine 17 (4): 500–503.

Groot, Vincent P., Georgios Gemenetzis, Alex B. Blair, Ding Ding, Ammar A. Javed, Richard A. Burkhart, Jun Yu, et al. 2018. “Implications of the Pattern of Disease Recurrence on Survival Following Pancreatectomy for Pancreatic Ductal Adenocarcinoma.” Annals of Surgical Oncology 25 (8): 2475–83.

Hermann, Chris D., Benjamin Schoeps, Celina Eckfeld, Enkhtsetseg Munkhbaatar, Lukas Kniep, Olga Prokopchuk, Nils Wirges, et al. 2021. “TIMP1 Expression Underlies Sex Disparity in Liver Metastasis and Survival in Pancreatic Cancer.” The Journal of Experimental Medicine 218 (11). https://doi.org/10.1084/jem.20210911.

Kim, Jaegil, David Kwiatkowski, David J. McConkey, Joshua J. Meeks, Samuel S. Freeman, Joaquim Bellmunt, Gad Getz, and Seth P. Lerner. 2019. “The Cancer Genome Atlas Expression Subtypes Stratify Response to Checkpoint Inhibition in Advanced Urothelial Cancer and Identify a Subset of Patients with High Survival Probability.” European Urology 75 (6): 961–64.

Kim, Jinkook, Eunjeong Ji, Kwangrok Jung, In Ho Jung, Jaewoo Park, Jong-Chan Lee, Jin Won Kim, Jin-Hyeok Hwang, and Jaihwan Kim. 2021. “Gender Differences in Patients with Metastatic Pancreatic Cancer Who Received FOLFIRINOX.” Journal of Personalized Medicine 11 (2). https://doi.org/10.3390/jpm11020083.

Kim, S-E, P. D. Cole, R. C. Cho, A. Ly, L. Ishiguro, K-J Sohn, R. Croxford, B. A. Kamen, and Y-I Kim. 2013. “^γ^-Glutamyl Hydrolase Modulation and Folate Influence Chemosensitivity of Cancer Cells to 5-Fluorouracil and Methotrexate.” British Journal of Cancer 109 (8): 2175–88.

Leibowitz, Benjamin D., Bonnie V. Dougherty, Joshua S. K. Bell, Joshuah Kapilivsky, Jackson Michuda, Andrew J. Sedgewick, Wesley A. Munson, et al. 2022. “Validation of Genomic and Transcriptomic Models of Homologous Recombination Deficiency in a Real-World Pan-Cancer Cohort.” BMC Cancer 22 (1): 587.

Long, Qi, Jianpeng Xu, Adeboye O. Osunkoya, Soma Sannigrahi, Brent A. Johnson, Wei Zhou, Theresa Gillespie, et al. 2014. “Global Transcriptome Analysis of Formalin-Fixed Prostate Cancer Specimens Identifies Biomarkers of Disease Recurrence.” Cancer Research 74 (12): 3228–37.

Lowery, Maeve A., Emmet J. Jordan, Olca Basturk, Ryan N. Ptashkin, Ahmet Zehir, Michael F. Berger, Tanisha Leach, et al. 2017. “Real-Time Genomic Profiling of Pancreatic Ductal Adenocarcinoma: Potential Actionability and Correlation with Clinical Phenotype.” Clinical Cancer Research. https://doi.org/10.1158/1078-0432.ccr-17-0899.

Ma, Nan, Lu Si, Meiling Yang, Meihua Li, and Zhiyi He. 2021. “A Highly Expressed mRNA Signature for Predicting Survival in Patients with Stage I/II Non-Small-Cell Lung Cancer after Operation.” Scientific Reports 11 (1): 5855.

Michuda, Jackson, Alessandra Breschi, Joshuah Kapilivsky, Kabir Manghnani, Calvin McCarter, Adam J. Hockenberry, Brittany Mineo, et al. 2022. “Validation of a Transcriptome-Based Assay for Classifying Cancers of Unknown Primary Origin.” bioRxiv. https://doi.org/10.1101/2022.05.06.22274683.

Moffitt, Richard A., Raoud Marayati, Elizabeth L. Flate, Keith E. Volmar, S. Gabriela Herrera Loeza, Katherine A. Hoadley, Naim U. Rashid, et al. 2015. “Virtual Microdissection Identifies Distinct Tumor- and Stroma-Specific Subtypes of Pancreatic Ductal Adenocarcinoma.” Nature Genetics. https://doi.org/10.1038/ng.3398.

Mueller, Sebastian, Thomas Engleitner, Roman Maresch, Magdalena Zukowska, Sebastian Lange, Thorsten Kaltenbacher, Björn Konukiewitz, et al. 2018. “Evolutionary Routes and KRAS Dosage Define Pancreatic Cancer Phenotypes.” Nature 554 (7690): 62–68.

Natale, Christopher A., Jinyang Li, Jason R. Pitarresi, Robert J. Norgard, Tzvete Dentchev, Brian C. Capell, John T. Seykora, Ben Z. Stanger, and Todd W. Ridky. 2020. “Pharmacologic Activation of the G Protein-Coupled Estrogen Receptor Inhibits Pancreatic Ductal Adenocarcinoma.” Cellular and Molecular Gastroenterology and Hepatology 10 (4): 868–80.e1.

O’Kane, Grainne M., Barbara T. Grünwald, Gun-Ho Jang, Mehdi Masoomian, Sarah Picardo, Robert C. Grant, Robert E. Denroche, et al. 2020. “GATA6 Expression Distinguishes Classical and Basal-like Subtypes in Advanced Pancreatic Cancer.” Clinical Cancer Research: An Official Journal of the American Association for Cancer Research 26 (18): 4901–10.

Orth, Michael, Philipp Metzger, Sabine Gerum, Julia Mayerle, Günter Schneider, Claus Belka, Maximilian Schnurr, and Kirsten Lauber. 2019. “Pancreatic Ductal Adenocarcinoma: Biological Hallmarks, Current Status, and Future Perspectives of Combined Modality Treatment Approaches.” Radiation Oncology 14 (1): 141.

Paik, Kwang Yeol, Seong Ho Choi, Jin Seok Heo, and Dong Wook Choi. 2012. “Analysis of Liver Metastasis after Resection for Pancreatic Ductal Adenocarcinoma.” World Journal of Gastrointestinal Oncology 4 (5): 109–14.

Park, Henry S., Shane Lloyd, Roy H. Decker, Lynn D. Wilson, and James B. Yu. 2012. “Overview of the Surveillance, Epidemiology, and End Results Database: Evolution, Data Variables, and Quality Assurance.” Current Problems in Cancer 36 (4): 183–90.

Peters, G. J., H. H. J. Backus, S. Freemantle, B. van Triest, G. Codacci-Pisanelli, C. L. van der Wilt, K. Smid, et al. 2002. “Induction of Thymidylate Synthase as a 5-Fluorouracil Resistance Mechanism.” Biochimica et Biophysica Acta 1587 (2-3): 194–205.

Pijnappel, Esther N., Melinda Schuurman, Anna D. Wagner, Judith de Vos-Geelen, Lydia G. M. van der Geest, Jan-Willem B. de Groot, Bas Groot Koerkamp, et al. 2022. “Sex, Gender and Age Differences in Treatment Allocation and Survival of Patients With Metastatic Pancreatic Cancer: A Nationwide Study.” Frontiers in Oncology 12 (March): 839779.

Pishvaian, Michael J., Edik M. Blais, Jonathan R. Brody, Emily Lyons, Patricia DeArbeloa, Andrew Hendifar, Sam Mikhail, et al. 2020. “Overall Survival in Patients with Pancreatic Cancer Receiving Matched Therapies Following Molecular Profiling: A Retrospective Analysis of the Know Your Tumor Registry Trial.” The Lancet Oncology 21 (4): 508–18.

Puleo, Francesco, Rémy Nicolle, Yuna Blum, Jérôme Cros, Laetitia Marisa, Pieter Demetter, Eric Quertinmont, et al. 2018. “Stratification of Pancreatic Ductal Adenocarcinomas Based on Tumor and Microenvironment Features.” Gastroenterology 155 (6): 1999–2013.e3.

Rashid, Naim U., Xianlu L. Peng, Chong Jin, Richard A. Moffitt, Keith E. Volmar, Brian A. Belt, Roheena Z. Panni, et al. 2020. “Purity Independent Subtyping of Tumors (PurIST), A Clinically Robust, Single-Sample Classifier for Tumor Subtyping in Pancreatic Cancer.” Clinical Cancer Research: An Official Journal of the American Association for Cancer Research 26 (1): 82–92.

Robinson, Dan R., Yi-Mi Wu, Robert J. Lonigro, Pankaj Vats, Erin Cobain, Jessica Everett, Xuhong Cao, et al. 2017. “Integrative Clinical Genomics of Metastatic Cancer.” Nature 548 (7667): 297–303.

Rodon, Jordi, Jean-Charles Soria, Raanan Berger, Wilson H. Miller, Eitan Rubin, Aleksandra Kugel, Apostolia Tsimberidou, et al. 2019. “Genomic and Transcriptomic Profiling Expands Precision Cancer Medicine: The WINTHER Trial.” Nature Medicine 25 (5): 751–58.

Sadahiro, Sotaro, Toshiyuki Suzuki, Akira Tanaka, Kazutake Okada, Akemi Kamijo, Hideki Nagase, and Junji Uchida. 2013. “Reduction in ^γ^-Glutamyl Hydrolase Expression Is Associated with Response to Uracil and Tegafur/leucovorin Chemotherapy in Patients with Colorectal Cancer.” Anticancer Research 33 (8): 3431–38.

Sadahiro, Sotaro, T. Suzuki, A. Tanaka, K. Okada, G. Saito, H. Miyakita, T. Ogimi, and H. Nagase. 2017. “Gene Expression Levels of Gamma-Glutamyl Hydrolase in Tumor Tissues May Be a Useful Biomarker for the Proper Use of S-1 and Tegafur-Uracil/leucovorin in Preoperative Chemoradiotherapy for Patients with Rectal Cancer.” Cancer Chemotherapy and Pharmacology 79 (6): 1077–85.

Saiki, Yuriko, Can Jiang, Masaki Ohmuraya, and Toru Furukawa. 2021. “Genetic Mutations of Pancreatic Cancer and Genetically Engineered Mouse Models.” Cancers 14 (1). https://doi.org/10.3390/cancers14010071.

Sakamoto, Etsuko, Sayaka Tsukioka, Shinji Oie, Takashi Kobunai, Hiroaki Tsujimoto, Kazuki Sakamoto, Yoshihiro Okayama, et al. 2008. “Folylpolyglutamate Synthase and Gamma-Glutamyl Hydrolase Regulate Leucovorin-Enhanced 5-Fluorouracil Anticancer Activity.” Biochemical and Biophysical Research Communications 365 (4): 801–7.

Sarantis, Panagiotis, Evangelos Koustas, Adriana Papadimitropoulou, Athanasios G. Papavassiliou, and Michalis V. Karamouzis. 2020. “Pancreatic Ductal Adenocarcinoma: Treatment Hurdles, Tumor Microenvironment and Immunotherapy.” World Journal of Gastrointestinal Oncology 12 (2): 173–81.

Surveillance, Epidemiology, and End Results (SEER) Program (www.seer.cancer.gov) SEER*Stat Database

Smit, V. T., A. J. Boot, A. M. Smits, G. J. Fleuren, C. J. Cornelisse, and J. L. Bos. 1988. “KRAS Codon 12 Mutations Occur Very Frequently in Pancreatic Adenocarcinomas.” Nucleic Acids Research 16 (16): 7773–82.

Sperti, C., C. Pasquali, A. Piccoli, and S. Pedrazzoli. 1997. “Recurrence after Resection for Ductal Adenocarcinoma of the Pancreas.” World Journal of Surgery 21 (2): 195–200.

Suenaga, Masaya, Tsutomu Fujii, Mitsuro Kanda, Hideki Takami, Norio Okumura, Yoshikuni Inokawa, Daisuke Kobayashi, et al. 2014. “Pattern of First Recurrent Lesions in Pancreatic Cancer: Hepatic Relapse Is Associated with Dismal Prognosis and Portal Vein Invasion.” Hepato-Gastroenterology 61 (134): 1756–61.

Sun, Li, Juan Li, Xiaomeng Li, Xuemei Yang, Shujun Zhang, Xue Wang, Nan Wang, Kanghong Xu, Xinquan Jiang, and Yi Zhang. 2021. “A Combined RNA Signature Predicts Recurrence Risk of Stage I-IIIA Lung Squamous Cell Carcinoma.” Frontiers in Genetics 12 (June): 676464.

Wallden, Brett, James Storhoff, Torsten Nielsen, Naeem Dowidar, Carl Schaper, Sean Ferree, Shuzhen Liu, et al. 2015. “Development and Verification of the PAM50-Based Prosigna Breast Cancer Gene Signature Assay.” BMC Medical Genomics 8 (August): 54.

Wenric, Stephane, James M. Davison, Yun E. Wang, Gregory M. Mayhew, Kirk Beebe, Hyunseok P. Kang, Michael V. Milburn, Vincent Chung, Tanios Bekaii-Saab, and Charles M. Perou. 2022. “Abstract A002: Purity Independent Subtyping of Tumor (PurIST): Real-World Data Validation of a Pancreatic Ductal Adenocarcinoma (PDAC) Gene Expression Classifier and Its Prognostic Implications.” Cancer Research 82 (22_Supplement): A002–A002.

Wise, Hannah C., and David B. Solit. 2019. “Precision Oncology: Three Small Steps Forward.” Cancer Cell.

Yachida, Shinichi, Catherine M. White, Yoshiki Naito, Yi Zhong, Jacqueline A. Brosnan, Anne M. Macgregor-Das, Richard A. Morgan, et al. 2012. “Clinical Significance of the Genetic Landscape of Pancreatic Cancer and Implications for Identification of Potential Long-Term Survivors.” Clinical Cancer Research: An Official Journal of the American Association for Cancer Research 18 (22): 6339–47.

