## Supplementary Materials for "Real-world data validation of the PurIST pancreatic ductal adenocarcinoma gene expression classifier and its prognostic implications"

**Title**

**Supplementary Materials**

**Results**

**Supplementary tables**

| **Characteristic** | **Basal, N = 19^1^** | **Classical, N = 81^1^** | **p^2^** |
| --- | --- | --- | --- |
| **Sex** |  |  | 0.10 |
| Female | 5 (26%) | 38 (47%) |  |
| Male | 14 (74%) | 43 (53%) |  |
| **Age at diagnosis** | 67 (60, 74) | 69 (59, 76) | 0.6 |
| **Stage** |  |  | 0.7 |
| I | 3 (16%) | 10 (12%) |  |
| II | 14 (74%) | 56 (69%) |  |
| III | 2 (11%) | 15 (19%) |  |
| **Histology** |  |  | >0.9 |
| Adenocarcinoma | 4 (21%) | 19 (23%) |  |
| Infiltrating duct carcinoma | 15 (79%) | 62 (77%) |  |
| ^1^n (%); Median (IQR) | | | |
| ^2^Pearson's Chi-squared test; Wilcoxon rank sum test; Fisher's exact test | | | |

**Supplementary table 1 -** Platform concordance cohort.

| **Site** | **Stage** | **Diluted purity** | **N Nocalls** | **N Samples** | **N Classical** | **N Basal** | **N Replicates** | **N Concordant** | **OPA** |
| --- | --- | --- | --- | --- | --- | --- | --- | --- | --- |
| Chicago | Repeatability | - | 1 | 8 | 5 | 3 | 23 | 23 | 1.00 |
| Chicago | Reproducibility | - | 1 | 11 | 6 | 5 | 33 | 33 | 1.00 |
| Chicago | LOD | 20% | 0 | 6 | 4 | 2 | 16 | 16 | 1.00 |
| Chicago | LOD | 30% | 0 | 6 | 4 | 2 | 15 | 15 | 1.00 |
| Chicago | LOD | 40% | 0 | 6 | 4 | 2 | 16 | 16 | 1.00 |
| Chicago | IIC | - | 0 | 9 | 4 | 5 | 27 | 27 | 1.00 |
| RTP | Repeatability | - | 1 | 11 | 6 | 5 | 31 | 31 | 1.00 |
| RTP | Reproducibility | - | 2 | 8 | 3 | 5 | 21 | 20 | 0.95 |
| RTP | LOD | 20% | 0 | 11 | 6 | 5 | 32 | 31 | 0.97 |
| RTP | LOD | 30% | 0 | 11 | 6 | 5 | 33 | 32 | 0.97 |
| RTP | LOD | 40% | 0 | 12 | 6 | 6 | 33 | 32 | 0.97 |
| RTP | IIC | - | 0 | 10 | 5 | 5 | 39 | 39 | 1.00 |

**Supplementary Table 2.** Analytical validation sample sizes and Overall Percent Agreement (OPA) between replicate subtypes and reference sample subtypes across the four stages of analytical validation at two Tempus Labs locations. LOD: Limit Of Detection; IIC: Inter-Instrument Concordance; RTP: Research Triangle Park, Durham, NC.

**Supplementary Figures**

**Supplementary Figure 1.** Platform concordance.

**SFig 1A.** Diagram of the platform concordance and retraining process.

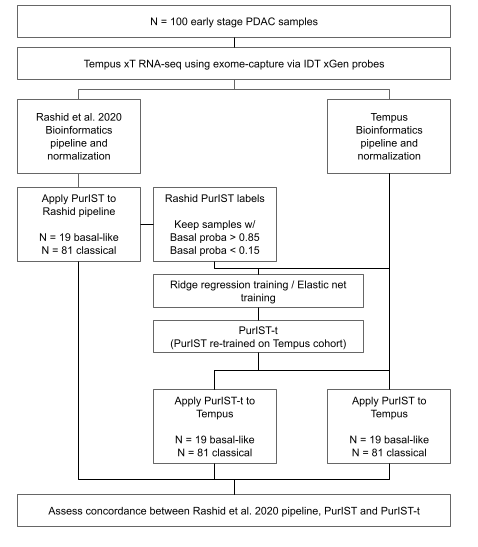

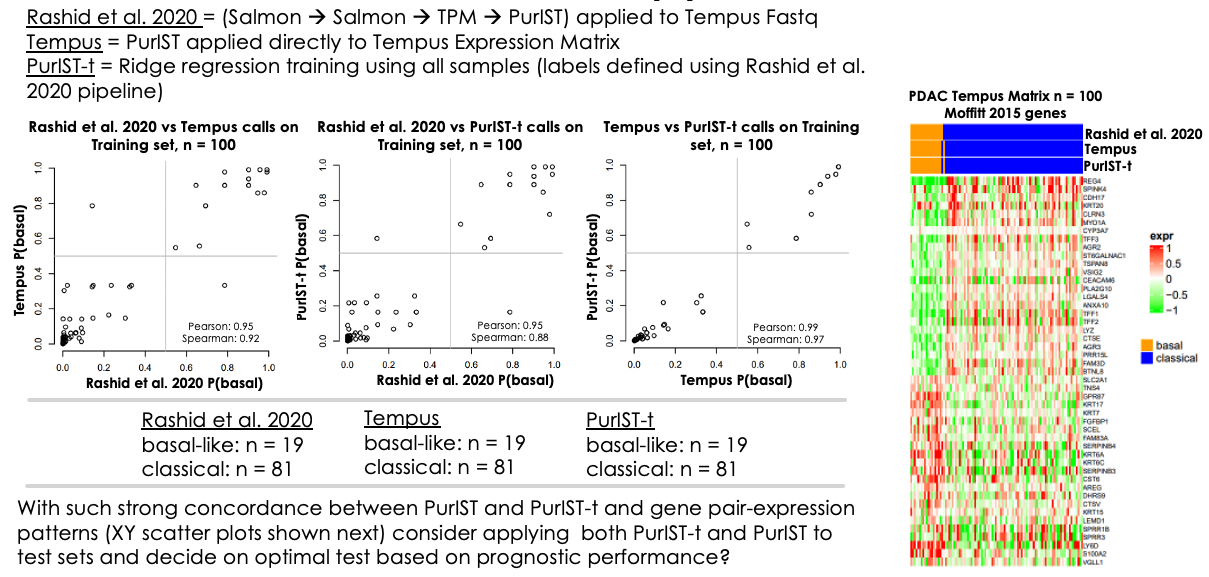

**SFig 1B.** Comparison of subtype calls obtained with the original PurIST model and the original bioinformatics pipeline; the original PurIST model and Tempus’ proprietary bioinformatics pipeline; the PurIST model re-trained on the platform concordance cohort.

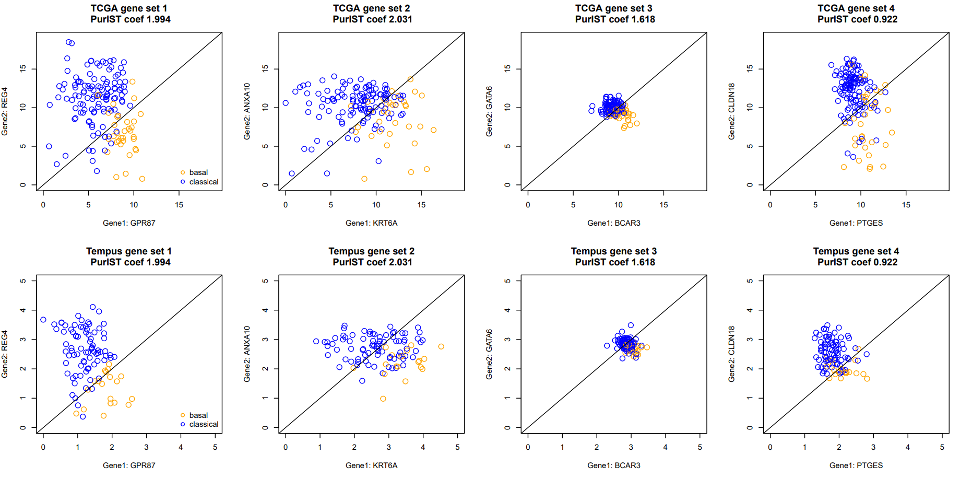

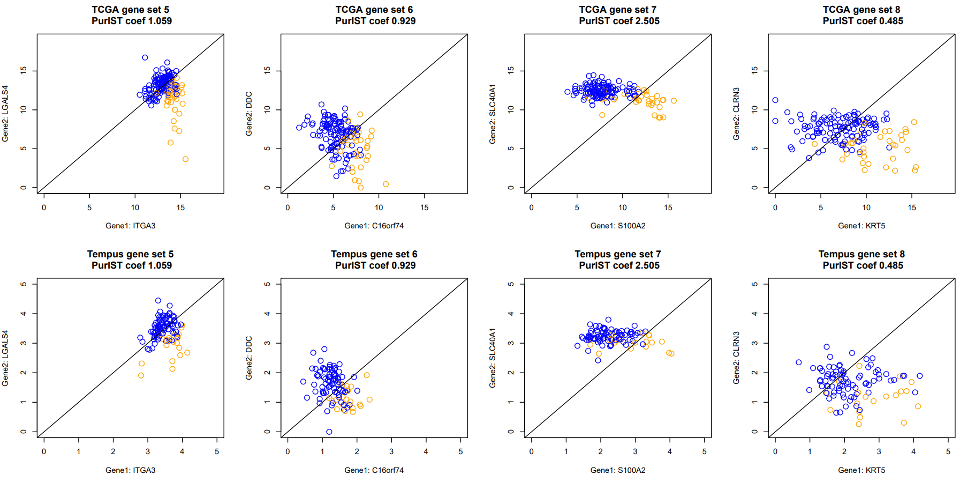

**SFig 1C.** XY scatter plots that display the distribution of normalized gene expression values of platform concordance cohort (rows 2 and 4) and TCGA PAAD cohort (rows 1 and 3) for the eight gene pairs in the PurIST algorithm. Each circle represents a sample and is colored based on its subtype.

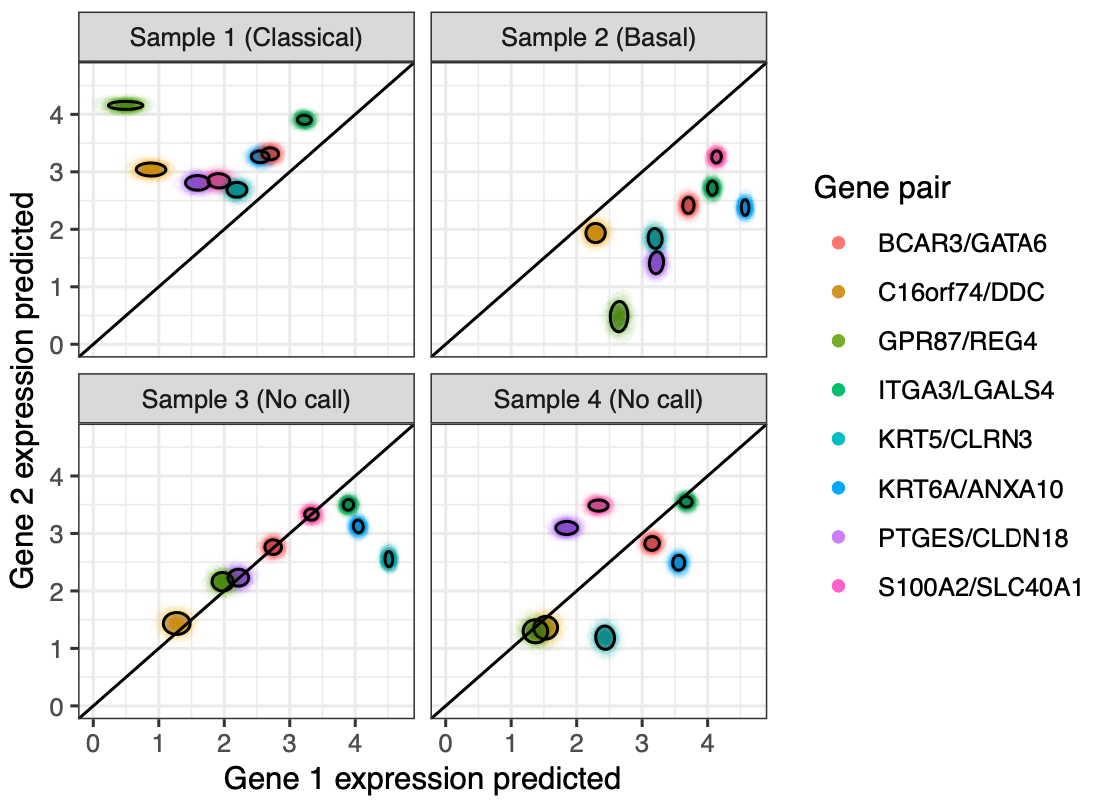

**Supplementary Figure 2.** Scatterplot of four sets of predicted PurISt gene expression values as a visual representation of the confidence scoring process for four selected samples. Each cloud of points includes N = 1000 simulated gene expression values for each gene pair. Black ellipses represent 0.85 confidence intervals, in line with the no-call threshold of 0.85 that was used in this study. Sample 1 is strongly predicted to be classical with a confidence score of 1.0; Sample 2 is strongly predicted to be basal with a confidence score of 1.0; Samples 3 and 4, with a mixture of pairwise expression values that fall on both sides of the 1:1 line, are flagged as no-calls because their respective confidence scores of 0.585 and 0.652 are below the 0.85 threshold.

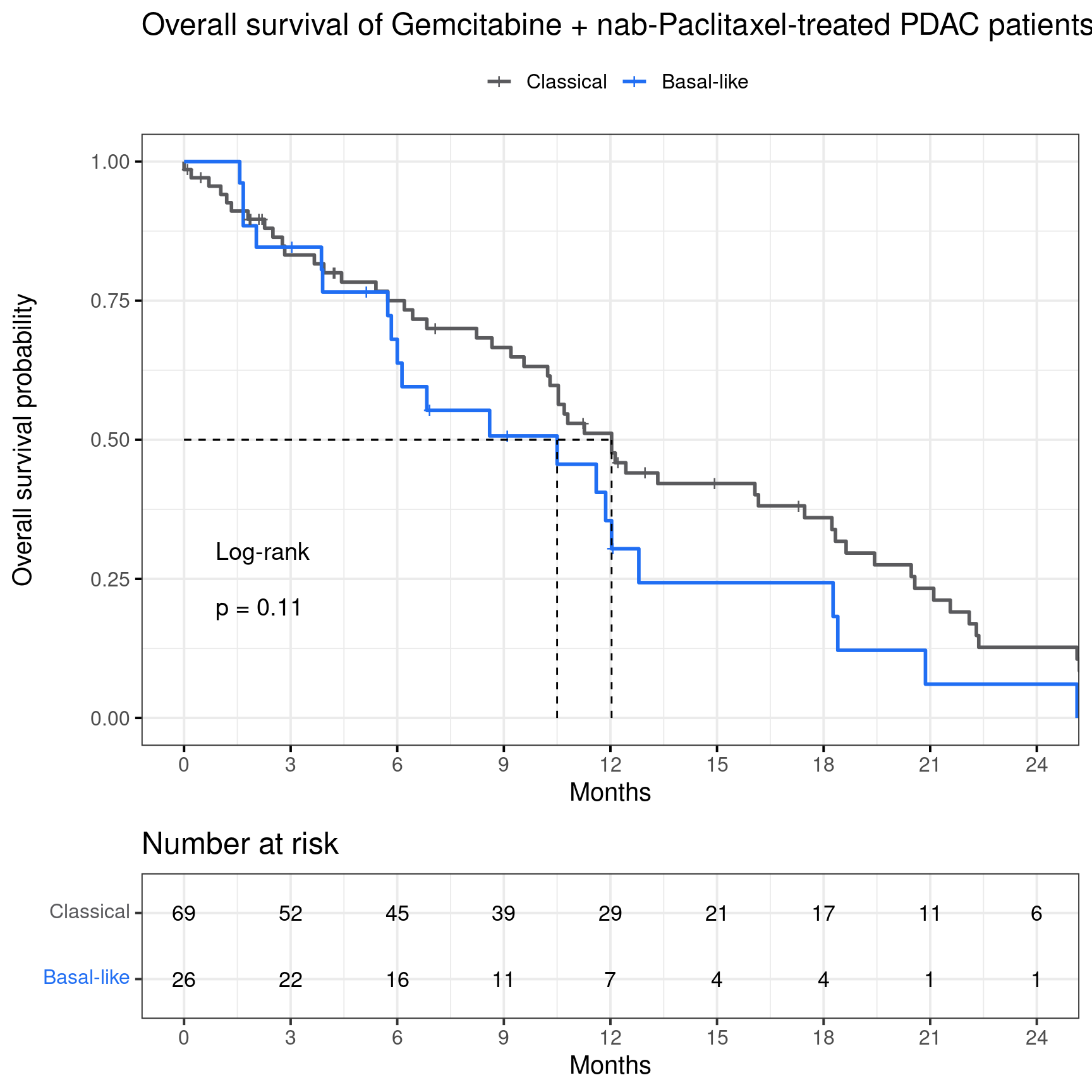

**Supplementary Figure 3**. Overall survival of the gemcitabine + nab-paclitaxel treatment group (n = 95).

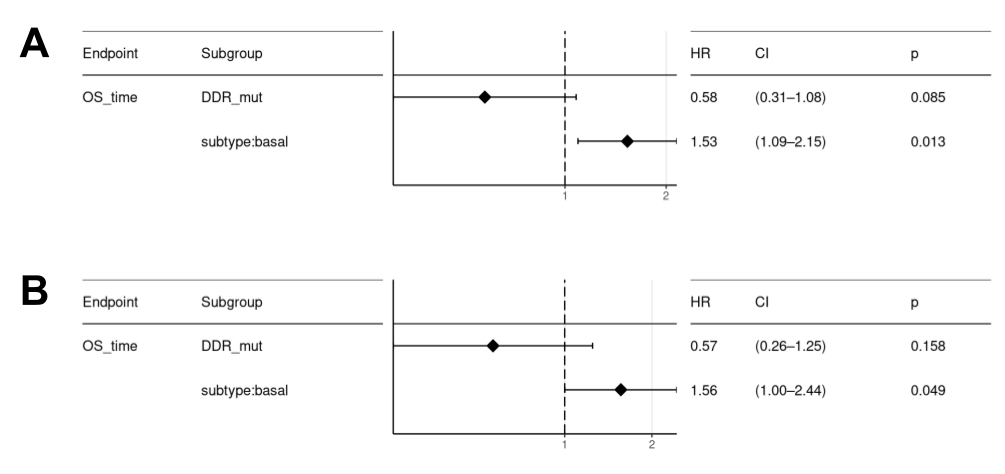

**Supplementary Figure 4.** Multivariate analysis including the presence of DDR mutations as covariate. **A**. Analysis on the full cohort (n = 232). **B**. Analysis on the FOLFIRINOX treatment group (n = 137)
